# Scaling Lassa Virus Dynamics within Anthropogenic Ecosystems (SCAPES) study: a mixed-methods observational cohort study of humans, rodents, and landscapes in Nigeria

**DOI:** 10.1101/2025.09.12.25335176

**Authors:** Sagan Friant, David Simons, Ottar Bjornstad, Rory Gibb, Christina Harden, Natalie Imirzian, Kate Jones, Abi Smith, Katharine Thompson, Martin Meremikwu, Lina Moses, David Redding

**Affiliations:** Department of Anthropology, The Pennsylvania State University, University Park, Pennsylvania USA; Huck Institutes of the Life Sciences, The Pennsylvania State University, University Park, Pennsylvania USA; Center for Infectious Disease Dynamics, The Pennsylvania State University, PA 16801; Department of Genetics, Evolution and Environment, University College London, London, UK; Science Department, Natural History Museum, Cromwell Road, London, SW7 5BD, United Kingdom; Institute of Tropical Diseases Research and Prevention, University of Calabar Teaching Hospital, Calabar, Nigeria; Tulane University School of Public Health and Tropical Medicine, New Orleans, LA, USA

## Abstract

**Introduction:** Zoonotic spillover driven by human activity remains a critical challenge. Research focusing on disease ecology or human–animal interactions provide detailed but often siloed local insights disconnected from global processes and policy. Risk maps are commonly used to inform decisions, yet their connection to local-scale drivers of transmission remains poorly understood. This study bridges these gaps through a cross-scale study of Lassa fever, a rodent-borne hemorrhagic fever of public health significance in West Africa and a WHO priority pathogen. We use a fine-scale quantitative and participatory modelling approach that explicitly integrates into broad-scale risk models to identify the patterns and processes that drive spillover within human-driven ecosystem at the animal-human interface. Lassa Fever epidemics are dominated by endemic and seasonal spillover of *Mammarenavirus lassaense* (LASV) from rodent reservoirs to humans within a rural context, positioning LASV as a uniquely tractable system in which to study spillover.

**Methods and Analysis:** We are currently conducting parallel observational studies of humans, rodents, and landscapes explicitly designed to feed into a cross-scale quantitative modelling framework. By integrating data on landcover, rodent dynamics, human behavior, and LASV infection, we examine how anthropogenic activities shape LASV ecology and human exposure. These data will inform spatial models of the human-rodent-LASV interface to predict key drivers of exposure risk. Finally, we integrate data from our empirical studies and emergent patterns from our interface model(s) into an existing broad-scale regression-based risk model. Our local-scale study and model predictions will clarify how zoonotic risk propagates from local to regional scales, providing evidence to inform disease management efforts for Lassa fever, and zoonotic spillover more generally.

**Ethics and dissemination:** Human subjects and animal research approvals were obtained from multiple ethical review boards, including the National Health Research Ethics Committee and National Veterinary Research Institute in Nigeria. The research protocol adheres to principles of ethical community-based health research including fostering collaboration, enhancing transparency, supporting capacity building, and disseminating results to communities and public health authorities.

**STRENGTHS AND LIMITATIONS OF THIS STUDY:** *Strengths:*

● Interdisciplinary approach: Integrates ecological, epidemiological, and anthropological methodologies for a comprehensive understanding of LASV dynamics.
● Predictive modeling: Utilizes modeling techniques to integrate diverse data sets, offering valuable insights for mechanistic understanding, forecasting, and intervention planning.
● Community engagement: Emphasizes ethical practices and active involvement of local communities, ensuring culturally sensitive research, guiding subsequent intervention design and enhancing data relevance.
● Geographical focus: Conducted in Nigeria within West Africa, a region critically affected by LASV, addressing a significant gap in current research and informing public health strategies.

*Limitations:*

● Complexity of integrating diverse data sets: Conceptual challenges in effectively integrating data from varied disciplines, which may require novel, integrative methodologies.
● Scaling local studies to regional interpretations of hotspots: Unknown capacity of geographically restricted, fine-scale data collections to make broader scale generalizations in risk modelling.
● Dependence on external factors: External factors such as political and economic stability, accessibility of remote areas, and public health policies might impact the study’s execution and outcomes.

## INTRODUCTION

Zoonotic spillover – the process by which a pathogen is transmitted from an animal to a human host – is a poorly understood phenomenon. Ecological and anthropological studies conducted at local scales are critical for understanding local-scale processes affecting spillover, but often lack the scope needed to explain global patterns of disease or to effectively inform policy decisions. Meanwhile, broad-scale (e.g. national or global-level) models forecasting spillover risk at policy relevant scales focus largely on understanding correlates risk without an understanding of underlying socio and ecological spillover mechanisms. Discontinuities between broad and local-scale models have limited the explanatory potential of either approach. In many contexts, broad-scale models are limited by lack of detailed socio-ecological data (e.g., spatially consistent information on human and reservoir behavior), and local-scale studies are not designed to explicitly fill these gaps. A unified approach is needed to understand how broad-scale patterns of risk manifest in local transmission dynamics, and how local scale processes, in turn, scale up to shape broader patterns of disease (Fig. 1).

**Figure 1.**
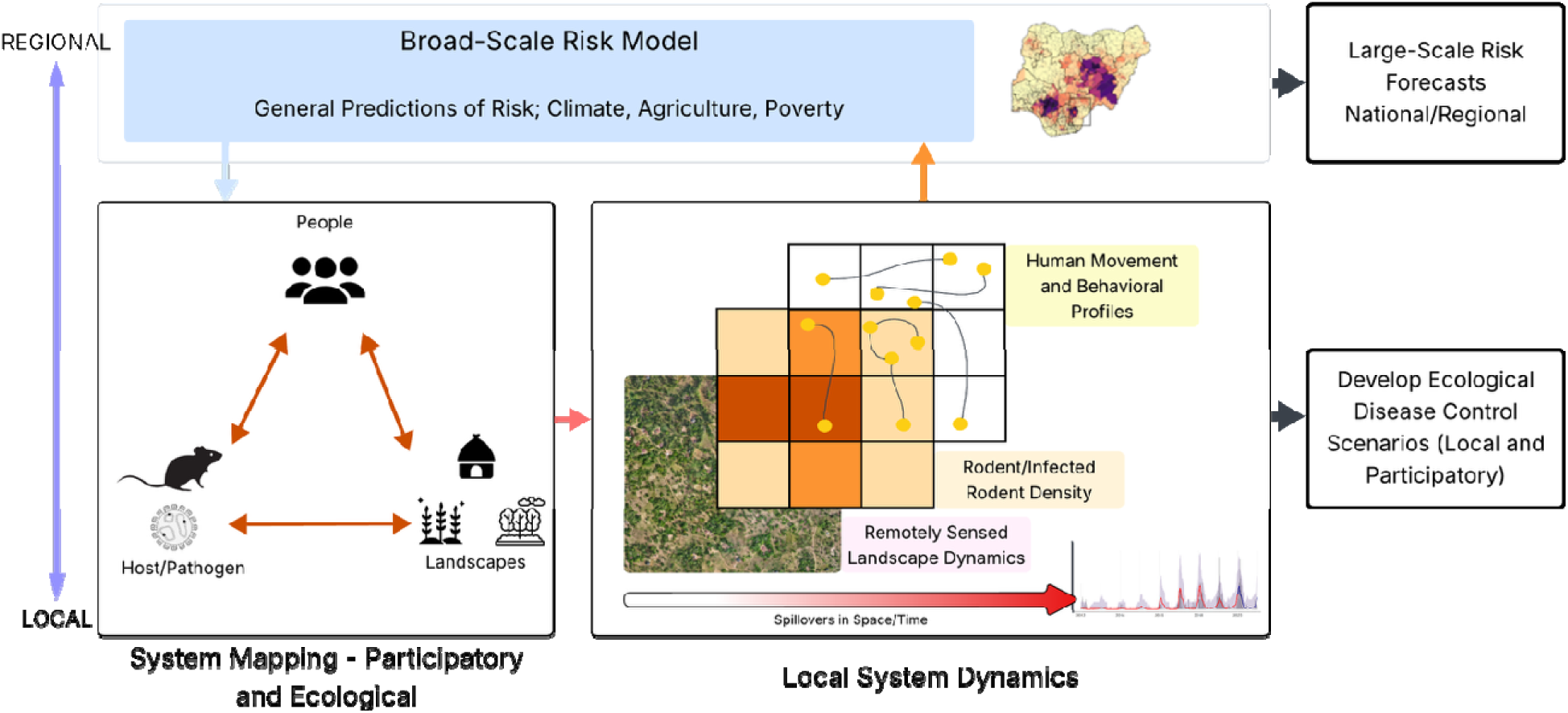
Cross-scale modelling framework. Local-scale data on human, rodent, and landscape factors are synthesized into socio-ecological motifs. These motifs inform interface models that identify the causal drivers of local transmission risk. The most robust drivers are then used to build a parsimonious, broad-scale predictive model. The final model is calibrated and validated using out-of-sample data from targeted field surveys in predicted high– and low-risk areas, ensuring the accuracy of the final risk maps.

Lassa fever is a rodent-borne hemorrhagic fever of public health significance in West Africa and a WHO priority pathogen (The Lancet Infectious Diseases 2018). Like many emerging zoonoses, Lassa virus (*Mammarenavirus lassaense*; LASV) spillover is characterized by multiple interacting species, spatial and temporal unpredictability, interacting biological and social factors, and difficulties in translating knowledge into management tools for public, agricultural, and ecosystem health (Gibb et al. 2017). The emergence and spread of LASV, as well as other zoonotic viruses, including hantaviruses, Nipah virus, Hendra virus, Ebola virus, and Marburg virus, is broadly associated with agricultural expansion and land use changes that alter the human-reservoir interface (Chua et al. 1999; Rose et al. 2001; Dearing and Dizney 2010; Wallace et al. 2014; Redding et al. 2016; Wallace et al. 2016; Kessler et al. 2018; Redding et al. 2021). Unlike other wildlife-origin zoonoses (e.g. Ebola, HIV, SARS-CoV1 & 2) which occur in sporadic outbreaks with subsequent human-to-human transmission chains that are spatially and temporally removed from spillover events (Ksiazek et al. 2003; Briand et al. 2014; Faria et al. 2016; Shereen et al. 2020), LASV spills over frequently, with zoonotic transmission accounting for the majority of human infections ranging from asymptomatic infection to acute hemorrhagic fever (Bausch et al. 2001; Lo Iacono et al. 2015; Andersen et al. 2015). Lassa fever therefore provides a uniquely tractable system to elucidate the drivers of spillover within human-driven ecosystems.

*Mastomys natalensis* (the multimammate rat) is the primary reservoir for LASV, although the role of other infected, sympatric small-mammal species in viral transmission and maintenance remains poorly understood (Fichet-Calvet et al. 2014; Olayemi et al. 2016; Agbonlahor et al. 2017; Olayemi et al. 2018). *Mastomys natalensis* is abundant and widespread across sub-Saharan Africa, dominating agricultural landscapes, while LASV infection within these rodents is limited to their West African range and heterogeneous within it (Simons et al. 2023; Gibb et al. 2017; Redding et al. 2016). Human infections may occur via direct contact with infected rodent bodily fluids or indirectly through contact with contaminated food, surfaces, or particles. Risk factors that alter the likelihood, route, and dose of infection are putatively linked to housing, food processing and storage, pest management, and hunting and consumption of rodents (WHO 2025); however, the exact transmission mechanism(s) and their underlying drivers remain poorly understood.

To understand LASV risk, we must consider the complex socio-ecological systems in which spillover occurs (Gibb et al. 2025; Friant et al., n.d.; Friant 2024) (Fig. 2). Human activities influence LASV spillover through both top-down forces (e.g., rodent hunting) and bottom-up pressures (e.g., land use and agricultural practices) forces that shape rodent population dynamics and human exposure. In LASV endemic regions, the primary reservoir, *M. natalensis*, is simultaneously a food source, an agricultural and household pest, and a key driver of zoonotic disease (Bonwitt et al. 2016; Douno et al. 2021; Stenseth et al. 2003). The social and ecological consequences of these interactions likely explain crucial epidemiological patterns, such as the seasonality of human outbreaks tied to crop cycles and rodent recruitment, and the clustering of infected rodent within specific households (Fichet-Calvet et al. 2007, 2008; Monadjem et al. 2011; Mariën et al. 2018, 2020). However, these fine-scale socioecological dynamics, together with the drivers of local transmission, are missing from current approaches to understanding and predicting risk. Understanding transmission therefore requires an integrated perspective as human-rodent-environment interactions are deeply embedded in local livelihoods and food systems.

**Figure 2.**
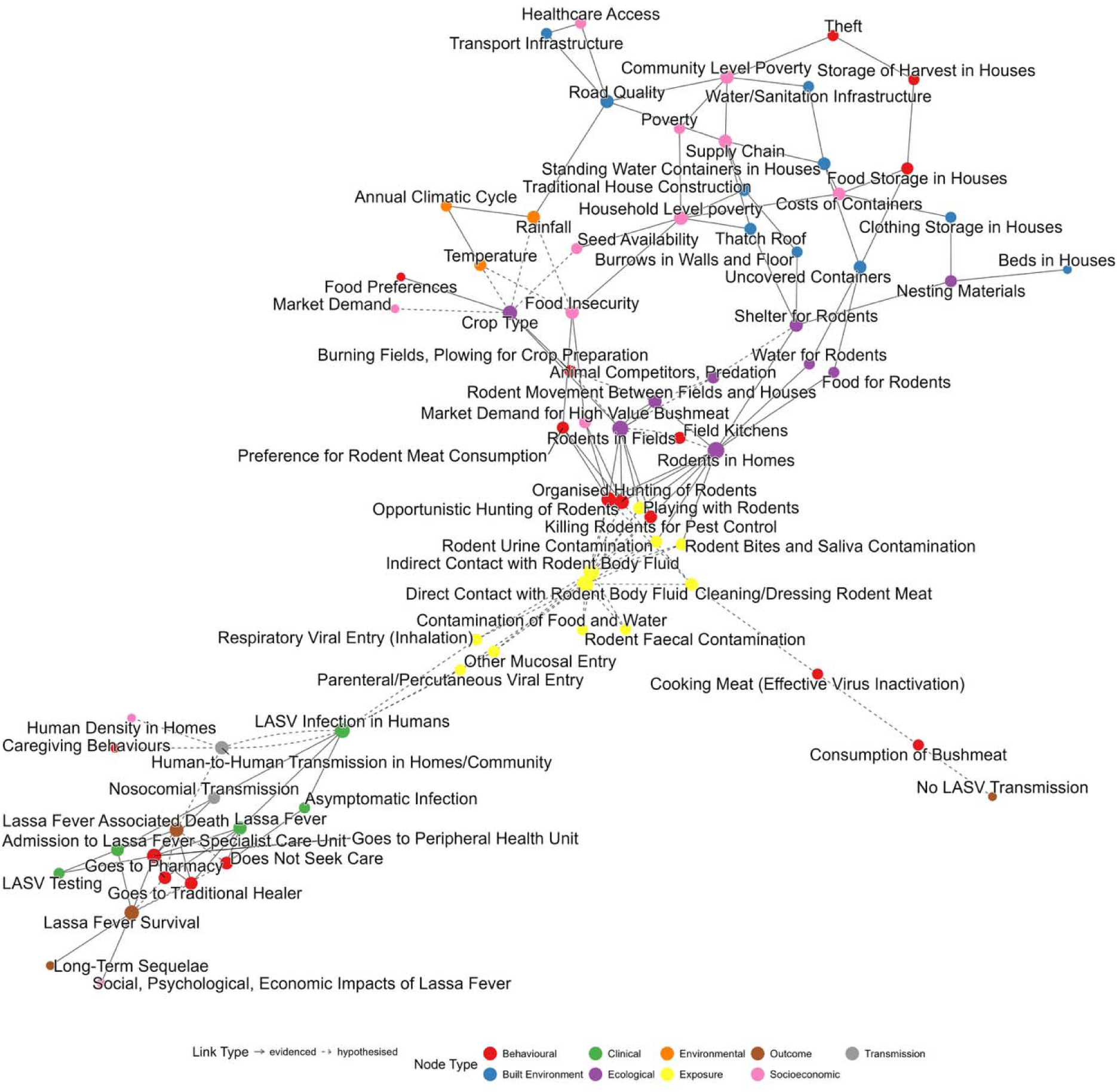
Conceptual model of the Lassa fever socio-ecological system. The network illustrates hypothesized causal pathways from upstream socio-ecological drivers to downstream clinical outcomes. Nodes are colored by conceptual link to Lassa fever and sized by their number of connections (degree centrality). Arrows indicate the direction of influence, while lines represent evidenced (solid) and hypothesized (dashed) links. For detailed annotations and references, see the interactive version of this model at the SCAPES project website.

Current approaches using broad-scale risk maps have focused on drivers like climate, land use, and poverty to predict where and when outbreaks may occur (Redding et al. 2021). While useful for high-level preparedness, these maps treat risk as an emergent property and are fundamentally limited because their inferential power is disconnected from the local-scale mechanisms that generate that risk. This scale mismatch, where policy is set broadly but interventions must be effective locally, hinders the development of targeted public health strategies. To truly understand and manage Lassa fever, and spillover infections more generally, we must bridge this gap by integrating knowledge of local processes across known axes of risk (e.g., rodent contact, climate, land-use and poverty) and throughout the range of LASV endemicity (i.e. from low-burden to high burden). Our project – **Sc**aling Lassa Virus Dynamics within **A**nthro**p**ogenic **E**co**s**ystems (SCAPES; <u>website</u>) – directly confronts this challenge by employing a framework (Fig. 1) to identify how emergent, broad-scale patterns of risk are generated from the collective fine-scale disease processes within human-driven environments.

### Aims and Objectives

The goal of this study is to contribute to the long-term reduction in Lassa fever burden in West Africa by: advancing our understanding of the complex dynamics of LASV spillover within human-modified ecosystems in West Africa, isolating those specific social and ecological contexts where spillover is most likely to occur, and using this improved systems understanding to develop more effective tools for forecasting zoonotic spillover. Stemming from this goal, we developed three interrelated aims:

1) Describe the interrelated dynamics of landscapes, humans, and rodents, that give rise to ‘hotspots’ of Lassa virus transmission risk using parallel field studies and integrated pattern-, process-, and participatory-based models to identify opportunities for disease control.
2) Predict risk of Lassa fever at larger spatial scales by integrating local-scale socio-ecological data into broad-scale risk models.
3) Communicate research findings to public health decision makers and local communities by co-developing intervention scenarios that foster evidence-based decision making and community engagement.

## METHODS AND ANALYSES

### Study Site and Population

The study will be conducted in selected sites within Nigeria, a country that reports the majority of Lassa fever cases but where disease risk is known to be markedly heterogeneous (Redding et al. 2021; Basinski et al. 2021). Our field studies will be centered around three focal sites in Nigeria that encompass variation across broad-scale axes of risk and extend from Lassa fever hotspots to the edge of its predicted distribution (Fig. 3A). Specifically, our study areas are within three states (Ebonyi, Benue, and Cross River) which vary in socioecological conditions expected to affect Lassa fever risk (Fig 3B).

**Figure 3.**
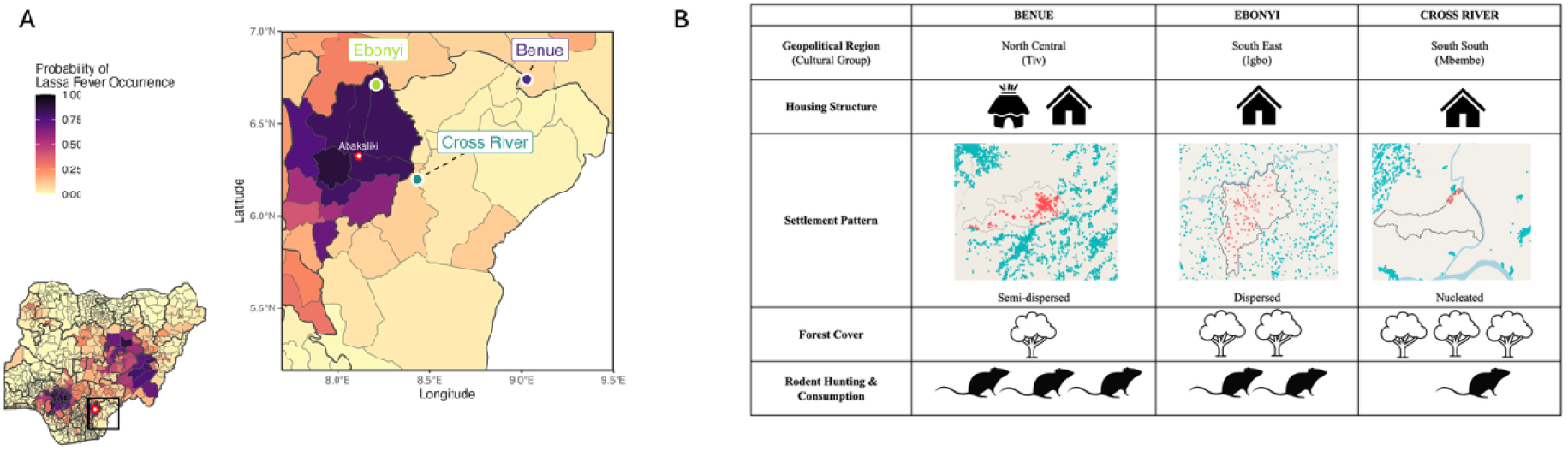
Study sites. The map shows the location of study villages within Nigeria and predicted probability of Lassa fever occurrence (A). Lassa fever occurrence was estimated from reported cases from 2018 to present. Socio-ecological conditions hypothesized to influence Lassa fever risk are expected to vary across sites (B). The three sites represent different geopolitical regions of Nigeria, dominant cultural groups, settlement patterns (e.g., spatially separated households surrounded by individual farmland vs. a single cluster of households with agricultural land spatially separated from villages), housing structures, land use practices, and cultural acceptance of small rodents as a food resource.

We identified specific study villages through a combination of cross-scale quantitative and qualitative assessments, integrating remotely sensed data with expert consultation and on-the-ground field visits to ensure local relevance and accuracy. Remotely sensed data were analyzed to select candidate field sites based on *a priori* quantitatively defined criteria: travel time (45 mins – 2 hours driving from our research base in Abakaliki, Ebonyi State), distance from major road (>1km), village size (100-500 households), settlement patterns (nucleated vs. dispersed houses) and landcover (predominantly agricultural, with <60% forest areas). An initial list of potential villages was obtained from OpenStreetMap and supplemented using the Geographic Names Server maintained by the National Geospatial Intelligence Agency (OpenStreetMap contributors. 2024; NGA and Esri 2024). Village size was estimated from the number of buildings using the Google OpenBuildings dataset (“Open Buildings” 2024). Landcover characteristics, primarily proportion tree cover, cropland, grassland, and shrubland, were estimated through the ESA WorldCover dataset (Zanaga et al. 2022). After identifying 426 villages within our region of interest, we narrowed down to 118 candidate sites based on the above quantitative criteria. We visually inspected sites on a satellite map to qualitatively assess whether they met our criteria. Villages were divided into a shortlist of three categories: nucleated villages with large distal fields, mixed housing and agriculture, and highly dispersed housing interspersed with fields. Shortlisted villages were assessed for feasibility, security, and reception to research through expert consultation at national, state, district, and community levels. Our quantitative criteria and expert consultation informed a hierarchy of villages based on accessibility or discordance with expected metrics. Study team members then conducted visits to candidate sites where they met with village heads and members from the community to assess fit (e.g., i.e., village size, land use types and major crops, and human-rodent interactions) as well as gather preliminary information on knowledge of rodents and Lassa fever. Ultimately, our focal sites were selected based on the receptiveness of the community to the research and research team to ensure that work could be conducted feasibly and safely.

### Study Design

Our field studies will incorporate epidemiological, anthropological, and ecological methodologies, tailored to capture the complex interplay between human activities, rodent reservoirs, and LASV transmission within human-driven landscapes (Fig. 4). Field activities will be conducted in three phases. Phase 1 includes a baseline cross-sectional serological survey for LASV antibodies, accompanied by household and individual questionnaires capturing demographics, structural characteristics, and rodent contact as well as spatial surveys and participatory mapping of landscape features and human activities. Phase 2 involves a two-year longitudinal study focused on anthropogenic landscapes, human movement and behavior, rodent ecology, and LASV surveillance within focal villages to collect parallel datasets across these domains. These local-scale field studies will inform a set of interface models to examine the spatial and temporal distribution of LASV in rodent populations, identify when and where humans overlap with infected rodents, and characterize the behaviors that drive exposure in relation to dynamic landscape processes. Data from empirical studies and emergent patterns from the interface models will be integrated back into existing broad-scale regression-based risk models to predict risk at larger spatial scales. Phase 3 consists of validation surveys at a broader spatial range to calibrate our predictive models.

**Figure 4.**
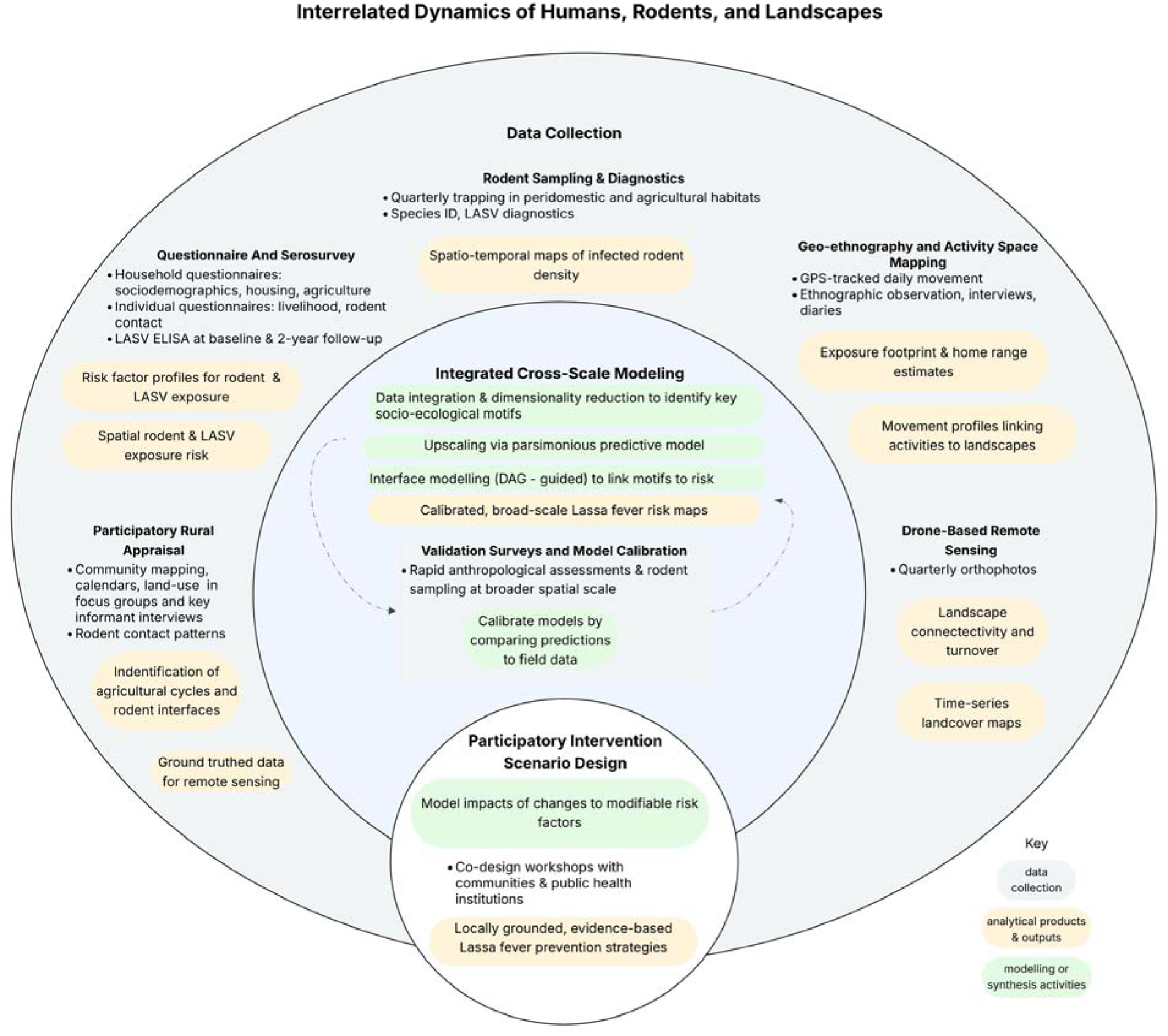
An integrated, cross-scale framework for the SCAPES project. The figure illustrates the project’s workflow, structured as three nested stages moving from broad data collection to focused application. The outer grey area represents data collection conducted in aim one to characterize the *Interrelated Dynamics of Humans, Rodents and Landscapes*, where five interlinked components gather empirical data. The orange shading reflects the primary outputs of this aim – the generation of analytical data products. These products feed into the blue ring, the *Integrated Cross-Scale Modeling Framework*, where the green shading signifies the focus on modeling activities. The central white circle represents the final aim, the *Intervention Scenario Design and Testing*. Here we synthesize all findings into actionable strategies grounded in the communities and policy landscapes of the project.

## AIM 1: Interrelated dynamics of humans, rodents, and landscapes

### Cross sectional baseline study

#### Participatory Rural Appraisal

We will use a collection of qualitative methods known as Participatory Rural Appraisal (PRA) to leverage local community knowledge of agricultural practices, land use, and human-rodent interfaces and ground our understanding of the socioecological system (Chambers 1994; Bernard 2011). Within each focal village, we will conduct focus group discussions and key informant interviews using specific PRA exercises, primarily community mapping and the development of seasonal calendars. These exercises will elicit detailed community knowledge on the spatial and temporal patterns of agricultural and non-agricultural land use, rodent ecology, and activities that mediate human-rodent contact. These qualitative data – such as community derived land cover classifications and timing of agricultural cycles (e.g., clearing, planting, harvesting) – will be used to define micro-zones of land use as well as ground-truth and annotate features on our remotely sensed maps. Local knowledge of human-rodent interfaces will inform the selection of key variables and the relationships between them, guiding both the causal structure and variable construction for our local-scale interface models.

#### Questionnaire and serosurvey

To identify the human behavioral and ecological factors that drive LASV transmission, we will conduct a mixed-methods, cross-sectional study across nine villages in our three study areas. Using a sequential explanatory design, qualitative data will expand upon the initial baseline quantitative data on human risk factors and LASV exposure. Together, these empirical data will inform the parameterization of local-scale interface models, calibration of broad-scale risk predictions, and support downstream intervention planning grounded in local realities.

A multi-module questionnaire will be administered to collect geocoded data on hypothesized risk factors. The instrument, informed by a systematic review of zoonotic disease literature, covers household sociodemographics, livelihood and agricultural practices, housing structure and location within the landscape, food storage methods, and the nature and frequency of human-rodent interactions. Specific modules will quantify food security, rodent-driven food loss, and document rodent off-take for consumption or pest control. Using systematic random sampling, we will enroll approximately 25% of households per village, targeting a total sample of 540 households for our quantitative questionnaire. Based on published and unpublished data, this sample size provides over 80% power (α=0.05) to detect statistically significant differences in seroprevalence between villages classified as high-risk (∼40% seroprevalence), medium-risk (∼20%), and low-risk (<5%) (Grant et al. 2023). Within each household, we will seek to enroll an adult male, an adult female, and a child (≥12 years) for individual questionnaires and serology, plus an additional child (<12 years) for serology only, yielding an estimated 1,620 completed individual questionnaires.

In the qualitative phase, we will conduct focus groups and in-depth interviews with men, women, children, as well as purposively selected individuals with specialist knowledge. These methods will deepen and contextualize our quantitative findings by capturing diverse perspectives on human-rodent interfaces. From these data, we will explore behavioral drivers of human–rodent contact and compare competing explanations of how people navigate trade-offs between minimizing risk and maximizing well-being within local food systems (e.g., hunting, crop protection, food storage). We will also identify perceived barriers and opportunities for rodent control that align with food security and disease prevention goals. Upon reaching saturation, these insights will be operationalized into emically grounded disease control scenarios and used to guide iterative feedback processes with local stakeholders.

As most LASV infections are subclinical, serological data are essential for accurately assessing cumulative exposure. Consenting participants will provide a finger-prick blood sample collected as a dried blood spot on filter paper, yielding an estimated 2,160 samples for analysis. These samples will be tested for LASV-specific IgG antibodies using a commercially available ELISA (Panadea® LASV IgG), providing a direct measure of past infection (Soubrier et al. 2022). To capture infection dynamics, we will collect replicate samples from the same individuals at the conclusion of Phase 2, allowing us to measure seroconversion and seroreversion rates and to identify risk factors for incident infections.

### Longitudinal parallel study

#### Drone-based remote sensing

To characterize the fine-scale landscape dynamics that structure human-rodent interactions, we will conduct quarterly drone-based remote sensing in each of the three focal villages. The spatial extent of these mapping campaigns will be guided by village boundaries established during the PRAs. Collected imagery will be processed into high-resolution orthomosaics, from which we will generate detailed, time-series land cover maps using a supervised computer vision model trained on PRA-informed, ground-truthed data (Burnett and Blaschke 2003). These locally validated land cover maps provide a foundational geospatial framework for integrating all project data streams. The ground-truthed maps will be used to characterize exposure surfaces – spatial datasets that capture important epidemiological information, such as building location and material, land use, crop types and stages, and the structural complexity of villages and agroecosystems (Tusting et al. 2019). We will then calculate spatial measures of connectivity between fields, houses and other key landscape features, as well as spatially explicit summaries of landscape complexity, turnover and evenness within focal buffers of 1m, 5m and 10m to serve as covariates in our interface models.

#### Geo-ethnography

To characterize the spatiotemporal dynamics of human behavior that mediate LASV exposure risk, we will conduct a longitudinal geo-ethnography (Matthews, Detwiler, and Burton 2005). This mixed-methods approach integrates quantitative movement data with quantitative and qualitative ethnographic data to understand not just where people go, but also what they do there that might lead to rodent contact and ultimately LASV exposure. A spatially stratified random subsample of approximately 30 households (∼90 individuals representing triads of men, women, and children from each household) across our three focal villages will be enrolled in this component and followed quarterly. Our data collection is layered: participants will carry a GPS tracking device for 7 days each quarter to map high-resolution movement patterns. These tracks will be contextualized using data from daily activity logs and 7-day recall questionnaires recording locations visited, major activities, and human-rodent interactions. To gain deeper insight, we will select a subset of participants, reflecting diverse livelihood activities and sociodemographic backgrounds, for direct participant observation. By documenting and geocoding the daily routines that shape the human-rodent interface, we will inform risk models by unifying individual-level data with landscape dynamics.

Our analytical framework is designed to fuse these data streams. First, we will process the GPS data to derive individual “exposure footprints” by calculating activity space measures (e.g., utilization, recursion, and intensity distributions) and estimating home ranges (Sherman et al. 2005; Burgoine et al. 2015; Nettles et al. 2022). These spatial footprints will then be systematically annotated using daily activity logs, weekly recall questionnaires, and our ethnographic field notes. This fusion allows us to quantify both the time spent in different land cover types (e.g., farms, forests, villages) and the frequency of risk-associated activities (e.g., crop processing and storage, hunting, cleaning) within them, enabling predictions about how specific movement patterns and activities relate to rodent exposure. We will then explore the social (e.g., gender, occupation) and environmental (e.g., seasonality, landscape composition) correlates of these patterns. The ultimate analytical goal is to derive a minimum set of socio-ecological movement profiles that capture key variation in space use, behavior, and social context across the study population. These profiles will serve as a critical intermediate product, linking landscape data, questionnaire responses, and LASV exposure within our final risk models.

#### Rodent sampling and diagnostics

To characterize the reservoir component of the transmission system, we will conduct longitudinal rodent sampling to estimate the abundance, distribution, and LASV infection in the small mammal community. Our design is spatially stratified by land cover types defined by our remote sensing and participatory mapping data, focusing on two key strata: 1) the agricultural and forest matrix, and 2) the peridomestic environment. In the agricultural/forest matrix, we will establish six 10×10 trapping grids, each with 50 traps (Pearson and Ruggiero 2003; Freeman et al. 2022). In the peridomestic environment, we will place approximately 400 traps within and around the households enrolled in our geo-ethnography cohort. We will use a mix of smaller Sherman and larger Tomahawk live traps to capture a wide diversity of rodent species. Trapping will be conducted for four consecutive nights each session, yielding a total trapping effort of approximately 86,400 trap-nights over the course of the study.

All captured animals will be processed in a field laboratory following appropriate biosafety procedures (Mills et al. 1995). For each individual, we will record demographic and morphological data and collect tissue samples (ear, blood, organs) for species identification (cytochrome-b PCR (Lecompte et al. 2005)), age estimation (morphometrics and DNA methylation (Arneson et al. 2022)), and LASV diagnostics.

Viral RNA will be detected using RT-PCR (RealStar® Lassa Virus RT-PCR Kit 2.0), and previous exposure will be assessed by testing for LASV-specific antibodies using the same ELISA protocol as for human samples (Soubrier et al. 2022). To analyze these data, we will use a hierarchical joint species distribution model (Doser, Finley, and Banerjee 2023). This framework will use the repeated trapping data to co-model the abundance and distribution of *M. natalensis* and other sympatric species across the landscape, while explicitly correcting for imperfect detection probability and accounting for interspecific interactions and environmental covariates. When integrated with parallel human and landscape data, rodent sampling will capture seasonal variation in rainfall, temperature, crop development, and livelihood activities influence rodent dynamics and distribution within anthropogenic landscapes. The primary output of this analysis will be predictive, spatially explicit maps of infected and non-infected rodent density for each sampling period, including uncertainty estimates, which are essential inputs for our interface models.

## AIM 2: Integrated Cross-Scale Modelling Framework

The central analytical goal of this project is to integrate our landscape, human, and rodent data to develop a cross-scale model of LASV transmission risk. This framework is designed to move beyond simple correlative risk models to identify and incorporate the key causal drivers of transmission into validated, predictive risk maps. The process involves three main stages: 1) data integration and dimensionality reduction; 2) interface modelling and scaling; and 3) out-of-sample model validation and calibration.

### Data integration and dimensionality reduction

To enable cross-disciplinary analyses, we will first integrate all quantitative data (e.g., human behaviour, landscape, and rodent metrics) and coded qualitative data into a unified analytic dataset with standardized variable formats, coding schemes, and measurement scales. To identify the most salient patterns driving variance in our dataset, we will then apply unsupervised machine learning techniques, such as non-negative matrix factorization. This approach will reduce the dataset’s dimensionality by extracting key latent variables or “motifs” (i.e., distinct socio-ecological profiles of behaviour or specific landscape configurations) that account for most of the variance (Nguyen and Holmes 2019). These derived motifs will serve as powerful, composite predictor variables in subsequent interface models.

### Interface modelling and scaling

Our core quantitative analysis will employ Bayesian mixed-effects models to integrate the various data streams. To explicitly model the human-rodent interface, we will construct joint models that link spatiotemporal landscape dynamics with layers of predicted infectious and non-infectious rodent density, and human movement profiles. These models will allow us to estimate the effects of individual-level variables (e.g., age, sex, livelihood), household-level factors (e.g., housing materials, wealth), and dynamic behavioural variables (e.g., time spent in specific agricultural areas) on an individual’s risk of LASV exposure in space and time. A key feature of our Bayesian approach will be the incorporation of informative priors derived from our qualitative research, ensuring our statistical models are grounded in the socio-cultural realities observed in the field (Choy, O’Leary, and Mengersen 2009).

All quantitative analyses will be guided by a formal causal inference framework. For each primary hypothesis, we will first develop a Directed Acyclic Graph based on an *a priori* conceptualisation of the socio-ecological system informed by the PRA and reviews of the published literature (Fig. 2). The purpose of each DAG is to make our hypothesized causal structures explicit, allowing us to distinguish between different types of covariates (Lipsky and Greenland 2022). This process is critical for developing statistical models that correctly estimate the causal effect of an exposure on an outcome by identifying: confounders (variables that are a common cause of both the exposure and outcome that must be adjusted for), colliders (variables that are a common effect of the exposure and another variable that must not be adjusted for), and mediators (variables that lie on the causal pathway). Following these principles, our final regression models will be specified to adjust for the minimal sufficient set of confounding variables identified in the DAG for each hypothesis, ensuring the validity of our causal estimates.

The predictive performance and robustness of our models will be initially assessed through a multi-pronged validation strategy (Poisot et al. 2025). Cross-validation techniques will assess model stability by applying our model to different spatial patches and time periods within our primary study sites. We will then compare model predictions with observed Lassa fever case data. Specifically, we will test the model’s capacity to reproduce the characteristic seasonal pulse of human infections and validate predictions against contemporaneous, district-level case surveillance data from the Nigeria Centre for Disease Control and observed seroconversion patterns in our cohort (Grimm and Railsback 2012). By testing these models at varying levels of data aggregation – from detailed local scales to broader regional scales – we can assess the scalability of our findings and the impact of spatial resolution on our understanding of LASV dynamics.

Following validation, we will develop a parsimonious predictive model of Lassa fever risk that can be applied across larger spatial and temporal scales. This model will incorporate the most stable and robust causal drivers identified through our earlier interface models, enabling the generation of broad-scale risk maps that are both empirically grounded and mechanistically informed. These maps will serve as tools for upscaling our findings, guiding further testing and refinement, and ultimately targeting interventions.

### Validation surveys and model calibration

To validate and calibrate our predictive models with independent, out-of-sample data, we will conduct targeted “light touch” field surveys across a broader spatial scale, spanning a gradient of high to low predicted Lassa fever risk. This phase directly addresses a fundamental bias in LASV eco-epidemiology – the historical concentration of surveillance in areas with known human disease. Using a risk stratified design, we will select 12 new villages based on model predictions, spanning predicted high-risk “hotspots” and low-risk “coldspots.” In each village, we will carry out rapid, concurrent data collection, including rodent trapping to assess *M. natalensis* presence and LASV prevalence as well as rapid anthropological assessments to characterize local human-rodent interfaces (Manderson and Aaby 1992). The primary analysis will compare observed field data to the model’s *a priori* predictions. This approach allows us to rigorously test model performance and assess which drivers best explain discrepancies. For example, if LASV in rodents is confined to predicted hotspots, it would suggest that reservoir ecology is the primary driver of risk. Conversely, widespread LASV presence regardless of predicted risk would point to variation in human-rodent interfaces as the key determinant. Findings from this validation phase will guide the development of a final, parsimonious predictive model with improved accuracy and ability to generate reliable, spatially explicit risk maps.

## AIM 3: Intervention scenario design and testing

To identify and communicate effective strategies for reducing the spread, spillover or human impacts of LASV infection, we will create locally derived control scenarios from our qualitative research with the *predicted outcomes from* our interface models. We will use these combined insights to explore counterfactual scenarios that examine how changes in local conditions might have influenced current patterns of LASV spillover. This approach will ensure that proposed interventions are both scientifically grounded and responsive to real work dynamics of the Lassa system. Scenarios will be informed by concerns about rodent damage to food resources, with the secondary goal of identifying “win-win” control methods that combat both disease control and food security.

We will engage local stakeholders in participatory scenario co-design to ensure intervention scenarios reflect local realities (Enfors et al. 2008; Li et al. 2014). Through facilitated focus groups, participants will review model outputs, interpret data, and subsequently assess feasibility and acceptability of potential control measures. This process will identify the components of the Lassa system that are most modifiable (e.g. rodent abundance, human behavior, or landscape dynamics). It will also ensure that proposed interventions are not only evidence based but aligned with the local priorities. Strategies that offer co-benefits, such as improved agricultural outcomes or food security, are expected to be more favorably received and therefore more effective. Finally, results will be shared via published reports at the national level and orally at community meetings at the local level. Feeback will be elicited through workshops with the NCDC and designated community liaisons at the local level to adapt project outcomes for maximum impact.

## PATIENT AND PUBLIC INVOLVEMENT STATEMENT

This study uses participatory methods that engage local communities at the forefront of global health challenges in the construction of knowledge and management strategies. Our research follows principles of ethical community-based health research including fostering collaboration, enhancing transparency, and supporting capacity building. As part of this effort, a community advisory board will be constructed in each community, with whom we will hold informational meetings and discussions explaining the study’s purpose, procedures, and potential benefits. This also provides the community with an opportunity to provide feedback on the research questions, implementations, and findings in real time. This not only enriches the data with local knowledge but also fosters community engagement and awareness, which are vital for the successful dissemination of results and implementation of any subsequent intervention strategies.

## CONTRIBUTIONS

The transmission of animal diseases to humans has significant consequences for public health and economies globally. When evaluating disease risks, policy decisions often rely on data visualization techniques, such as risk maps. However, there is a limited understanding of how broad-scale risk maps relate to local-scale processes that affect disease transmission from animals to humans. For example, risk maps are often missing vital information on how broad associations between disease risk and factors such as climate, land-use, and poverty are driven by interactions between humans, animals, and the environment that affect probability of human infection. On the other hand, localized studies revealing fine-scale disease processes are often poorly situated to inform projections of risk at broader spatial scales. This project bridges this gap through a study of Lassa fever, a rodent-borne hemorrhagic fever of public health significance in West Africa and a global health priority.

Lassa fever outbreaks are a yearly occurrence in Nigeria. There is currently no vaccine, and prevention of Lassa fever is dependent on early diagnosis and managing the human-reservoir interface. However, effective management of this interface is hampered by a limited understanding of the nature and extent of human-rodent interactions across West Africa. This knowledge gap is compounded by a historical sampling bias, as LASV surveillance has largely been concentrated in recognized ‘hotspots’.

Consequently, our ability to forecast risk for LASV and other endemic rodent-borne infectious diseases outside of these ‘known knowns’, and to develop socially and ecologically based interventions to prevent spillover remains limited. A major strength of this study is that our sampling strategy is explicitly designed to address this limitation, providing a more grounded understanding of spatial variation in risk.

Field studies within SCAPES sample across known broad-scale drivers of Lassa fever risk to understand how local-scale processes vary across scale (e.g., how reservoir population dynamics are impacted by human land-use, or how poverty translates to high-risk human behavior etc.). The outcomes of this research are expected to provide critical insights into patterns and processes of LASV spillover, contribute to global understanding of the virus, and inform targeted intervention strategies to mitigate the impact of LASV in West Africa. Understanding these processes within human-driven environments is of paramount importance, given the rapid and unprecedented rate at which the natural world becomes human-dominated and the associated rise of emerging infectious diseases at the human-animal interface. The COVID-19 pandemic has been a stark reminder of our failure to understand these systems fully and make reasonable predictions of zoonotic spillover. Further work in this area will improve our ability to predict and ultimately minimize spillover risk in human populations globally.

## ETHICS AND DISSEMINATION

Human subjects research has been approved by the National Health Research Ethics Committee in Nigeria for Human Subjects Research (NHREC/01/01/2007-17/07/2023; 8/24/2023), relevant State Ministries of Health, and the Pennsylvania State University Institutional Review Board (STUDY00019989). Animal research has been approved by the Nigeria National Veterinary Research Institute Animal Ethics Committee (AEC/03/168/24) and Tulane IACUC (Approval 2492).

The dissemination of our study findings is planned to occur through multiple channels, targeting academic, public health, and participant communities in order to maximize the impact and relevance of the study’s findings. Academic findings will be published in peer-reviewed journals and presented at international conferences, contributing to the broader scientific discourse on LASV, disease ecology, and public health. Our findings will also be disseminated within the communities involved in the study. Plans for dissemination are included as part of our participatory research, through our participatory scenario planning protocols. Other workshops and community meetings will be organized to ensure that the knowledge generated is made accessible to those most affected by LASV. We will also disseminate findings to public health authorities at the community, state and national levels through reports, workshops, and meetings. In addition to our project findings, this study also contributes to science and public health via the production of a suite of tools and robust data products for scientists adopting similar approaches and decision makers for improved epidemic preparedness. To ensure these tools are useable and useful to public health agencies, we will host feedback and training workshops with the Nigeria Centre for Disease Control. Thus, our proposed research has both theoretical and practical applications beyond our study system, not only for other disease systems, but also other fields of inquiry.

## AUTHORS CONTRIBUTIONS

SF, LM, DR and RG conceived of the study. All authors contributed to the development of the study design and protocol. SF, DS, and DR drafted the manuscript, and CH, AS, NI, and KT edited the manuscript.

## Data Availability

No data are included in this protocol manuscript

